# A Potential Pitfall in Detection of Diabetes Mellitus Non-invasively Using the Second-Derivative of Photoplethysmography: Coexisting Hypertension May Blunt the Distinctive Difference in Early Negative to Initial Positive Wave Ratio (b/a) Between People With and Without Diabetes

**DOI:** 10.1101/2022.12.17.22283608

**Authors:** Ahmet Tas, Yaren Alan, Ilke Kara, Abdullah Savas, Muhammed Ikbal Bayhan, Diren Ekici, Zeynep Atay, Fatih Sezer, Cagla Kitapli

## Abstract

**Background:** Digital biomarkers have attracted increasing attention as possible non-invasive diagnostic tools for frequent cardiovascular pathologies. Alterations of features extracted from the derivatives and raw signal of photoplethysmography have been found in the presence of cardiovascular risk factors including but not limited to diabetes, hypertension, arterial stiffness, and endothelial dysfunction. In parallel, recent studies have shown promising results regarding the utilization of PPG as a non-invasive diagnostic tool for diabetes. Whether the presence of hypertension impedes the classification due to mutual endothelial insult resulting in peripheral hemodynamic perturbations remains unknown.

**Methods:** The previously described ratio between the amplitudes of early-negative(b) and initial-positive(a) waves (b/a) was derived from the second derivative of the PPG signal. Patients were classified according to diabetes mellitus type 2 and hypertension. Standard statistical tests were used to compare the means between unpaired subgroups.

**Results:** Final analysis included 132 patients. Compared to healthy control group DM2(-)HT(-) (-0.263 ± 0.115), both DM2(+)HT(-) (-0.361 ± 0.122, p<0.001, *d*= 0.83) and HT(+)DM2(-) (-0.319 ± 0.127, p:0.033, *d*= 0.48) groups had significantly lower b/a values. However, subgroups with one of two conditions had statistically indifferent mean b/a values (DM2(+)HT(-) (-0.361 ± 0.122) vs HT(+)DM2(-) (-0.319 ± 0.127, p:0.212, *d*:0.32). Moreover, significant difference in b/a between DM2 and non-DM2 subgroups of non-HT group (p<0.001) disappeared in subgroups with HT(p:0.665, *d*:0.16).

**Conclusion:** The utilization of the second-derivative of PPG for the detection of diabetes non-invasively may be impeded in patients with cardiovascular comorbidities, especially in the hypertensive population since both HT and DM2 induce parallel b/a ratio alterations due to the mutual undesired perturbations in the cardiovascular system, affecting peripheral flow dynamics.

## Background

The urge to develop non-invasive screening methods for common cardiovascular diseases continues. Photoplethysmography (PPG) is a low-cost, practical and widely-available non-invasive tool measuring the volumetric changes in blood flow[1]. Waveforms extracted from the second-derivative of the PPG (SD-PPG) signal, indicating the acceleration characteristics of flow velocity, have been shown to reflect endothelial function[2], hypertension(HT) [3], glucose intolerance[4], dyslipidemia, impaired fasting glucose [3], coronary heart disease risk, age, arterial stiffness, atherosclerosis [5] and diabetes mellitus(DM)[3, 6]. Moreover, multiple studies evaluating the PPG as a potential noninvasive tool to detect blood glucose levels as well as DM have provided encouraging results [7]. The b/a ratio, representing the ratio between the amplitudes of early-negative(b) and initial-positive(a) waves in SD-PPG, was shown to be associated with DM, impaired fasting glucose and impaired glucose challenge test response [3, 4, 6]. In parallel, ‘b/a’ was a successful predictor of DM in a recent classifier model [6]. However, the same ratio is also related to endothelial function [2], arterial stiffness [5], and ultimately hypertension [3]. Considering the frequent coexistence of HT and DM2, whether the b/a remains useful in the detection of DM2 in the hypertensive population, is an important question to answer. We hypothesize that, due to the mutual undesired endothelial and hemodynamic impacts of HT and DM2, the previously reported difference of b/a between DM2 and non-DM2 groups may disappear in the presence of HT.

## Methods

To test our hypothesis, we used an open dataset containing PPG signal and information about DM2, HT, age and sex[8]. Subjects with cerebrovascular pathologies(n=46) were excluded. The second derivative of the PPG signal was computed using MATLAB (MathWorks,USA) and the peaks of the a and b waves were manually extracted and averaged (Figure 1A). For dichotomic assessment, patients with ‘stage 1-2 hypertension’ were labeled as ‘HT’ whereas patients originally named as ‘normotensive-prehypertensive’ were labeled as ‘non-HT’.

**Fig 1.**
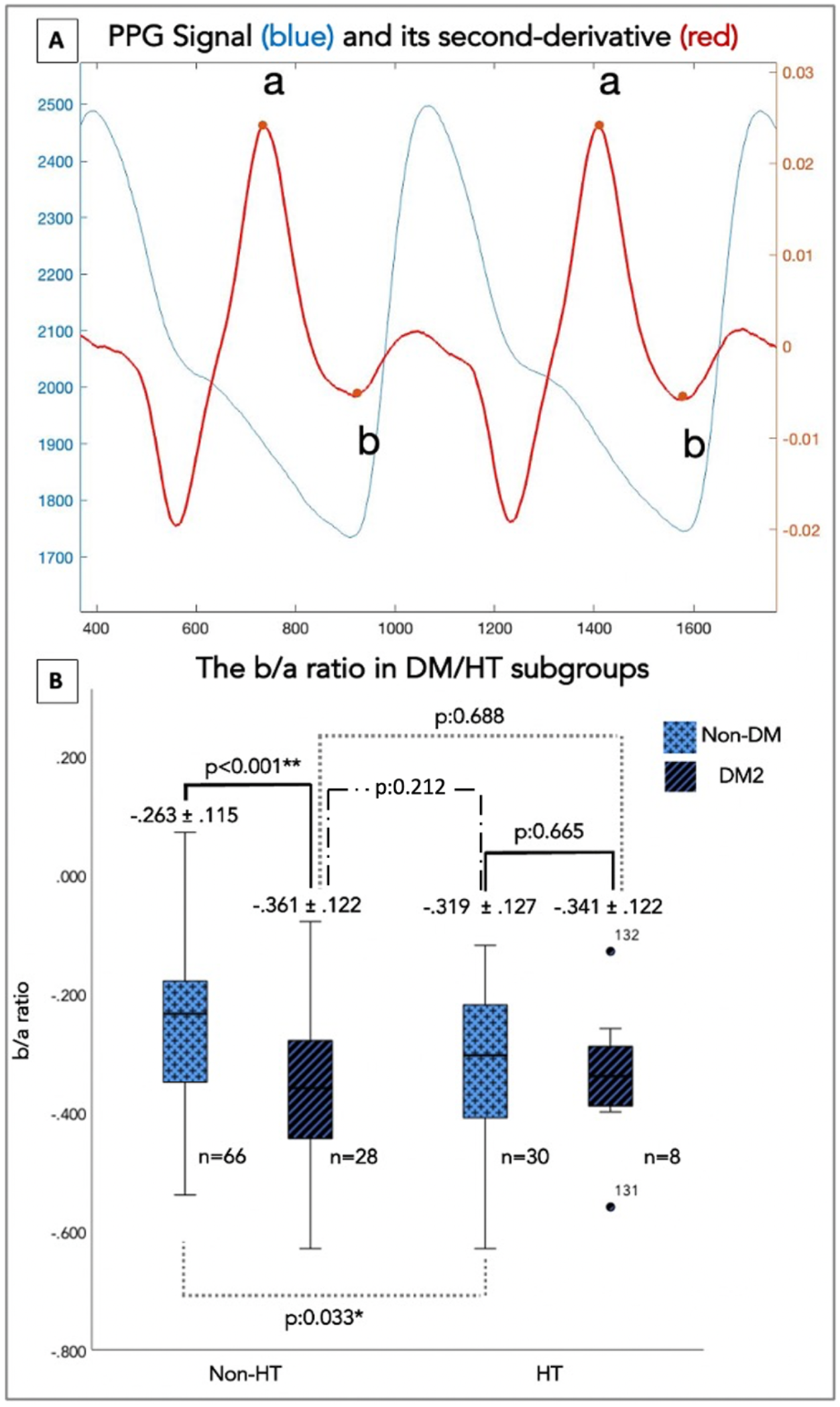
Example of PPG Waveforms (A) and Comparison of b/a between study subgroups(B). Compared to healthy control group [DM2(-)HT(-): -0.263 ± 0.115)], both [DM2(+)HT(-) -0.361 ± 0.122,, p<0.001, *d*= 0.83] and [HT(+)DM2(-) -0.319 ± 0.127, p:0.033, *d*= 0.48] groups had significantly lower b/a values. However, subgroups with one of two conditions have statistically indifferent mean b/a values (DM2(+)HT(-) -0.361 ± 0.122 vs HT(+)DM2(-) -0.319 ± 0.127, p:0.212, *d*:0.32). Moreover, significant difference in b/a between DM2 and non-DM2 subgroups of non-HT group (p<0.001) disappears in subgroups with HT(p:0.665, *d*:0.16).

Since the mean ages, which is known to affect the b/a ratio, were different between DM2(-/+) non-HT subgroups, we formed an age-matched control group of HT(-)DM22(-) prior to analysis, by including only the subjects whose ages were within the mean ± SD age range of HT(-)DM(+). In the HT group, mean ages were indifferent between DM2 and non-DM2 subgroups, thus no adjustments were made. Data was analyzed offline using SPSS (IBM,USA). Continuous variables were expressed as mean ± SD. Normality was assessed with the Shapiro-Wilk test. Means were compared using the Student’s t and Mann-Whitney U tests accordingly. Cohen’s *d* was used to interpret effect sizes. P<0.05 was considered statistically significant.

The present study was performed on a previously published free-use open-dataset containing data from deidentified patients [8]. Hence, ethical approval was not sought.

## Results

Figure 1B demonstrates the main findings. The final analysis included 132 patients .Three patients with inadequate signal quality were excluded. Fifty-one percent of patients were male. In the non-HT group, there were twenty-eight patients with DM2 and sixty-six patients without DM2, whereas in the HT group, there were eight patients with DM2 and thirty patients without DM2. Mean ages were indifferent between non-hypertensive patients with/without DM2 (57.3 ± 12.1 vs 53.8 ± 9.2, p:0.228) and between hypertensive patients with/without DM2 (63.4 ± 10.2 vs 60.9 ± 10.7, p:0.555).

In the non-HT group, patients with DM2 had significantly lower b/a ratio compared to those without DM2(-0.361 ± 0.122 vs -0.263 ± 0.115, p<0.001, *d*= 0.83). In parallel, in the non-DM2 group, patients with hypertension had significantly lower b/a values compared to those without HT (-0.319 ± 0.127 vs -0.263 ± 0.115,p:0.033, *d*:0.48), however, in the HT group the mean b/a ratio was indifferent between patients with and without DM2(-0.341±0.122 vs -0.319 ± 0.127, p:0.665, *d*:0.16). Likewise, mean the b/a ratios were indifferent between DM2(+)HT(-) and HT(+)DM2(-) groups (p:0.212, *d*:0.32) meaning that subgroups with one of the two studied cardiovascular risk factors had statistically inseparable mean b/a ratios.

## Discussion

The present study has two major findings. Firstly, the patients with DM2 or HT had significantly lower b/a ratios compared to healthy subjects (DM(-)HT(-)) which is in general consistent with the literature [7].

Importantly, our findings should be dealt with caution when comparing them to those of previous studies, since the reported (b/a) ratios may be absolute values as in [9]. Secondly, patients who had either DM2 or HT had similar b/a ratios, which can be justified by a miscellanea of preceding studies showing the relationships between the b/a ratio, and arterial stiffness[5], hypertension[10], impaired glucose tolerance[11] and fasting glucose as well as dyslipidemia[12]. Moreover, although the difference between means was statistically insignificant between HT(+)DM2(+) and HT(+)DM2(-) subgroups, the small sample size and numerical tendency towards lower values in the overlap group may encourage further research with the focus of overlapping cardiovascular risk factors. While the former finding is confirmatory in its nature, the latter result uniquely indicates a potential pitfall in detection of diabetes non-invasively, using digital biomarkers with the second-derivative of photoplethysmography.

## Conclusion

The utilization of the second-derivative of PPG for the detection of diabetes non-invasively may be impeded in patients with cardiovascular comorbidities, especially in the hypertensive population since both HT and DM2 induce parallel b/a ratio alterations due to the mutual undesired perturbations in the cardiovascular system, affecting peripheral flow dynamics.

## Data Availability

All data produced in the present study are available upon reasonable request to the authors.

## Abbreviations

DM: Diabetes Mellitus
HT: Hypertension
PPG: Photoplethysmography
SDPPG: Second-derivative of Photoplethysmography

